# Syphilis self-testing to expand test uptake among men who have sex with men: a theoretically informed mixed methods study in Zimbabwe

**DOI:** 10.1101/2020.11.30.20240788

**Authors:** Clarisse Sri-Pathmanathan, Definate Nhamo, Takudzwa Mamvuto, Gwendoline Chapwanya, Fern Terris-Prestholt, Imelda Mahaka, Michael Marks, Joseph D. Tucker

## Abstract

**Objectives:** Self-testing for STIs such as HIV and syphilis may empower sexual minorities and expand uptake of STI testing. While much is known about HIV self-testing (HIVST), less is known about syphilis self-testing, particularly in low-income settings. The objective of this study is to determine context-specific facilitators and barriers for self-testing and to assess the usability of syphilis self-testing in Zimbabwe among men who have sex with men (MSM).

**Methods:** This mixed methods study was conducted in Harare as part of a larger syphilis self-testing trial. The study included in-depth interviews (phase one) followed by usability testing and a second interview (phase two). In-depth interviews were conducted with MSM and key informants prior to syphilis self-testing. The same MSM then used the syphilis self-test, quantitatively assessed its usability and participated in a second in-depth interview. Phase one data was analysed using a thematic approach, guided by an adapted Social Ecological Model conceptual framework. Phase two interviews were analysed using Rapid Assessment Procedure qualitative methodology, and usability was assessed using a pre-established index, adapted from existing HIVST evaluation scales.

**Results:** Twenty MSM and 10 key informants were recruited for phase one in-depth interviews and 16 of these MSM participated in phase two by completing a syphilis self-test kit. Facilitating factors for self-testing included the potential for increased privacy, convenience, autonomy and avoidance of social and healthcare provider stigma. Barriers included the fear to test and uncertainty about linkage to care and treatment. Data from the usability index suggested high usability (89.6% on a 0-100 scale) among the men who received the self-test.

**Conclusions:** MSM in Zimbabwe were willing to use syphilis self-test kits and many of the barriers and facilitators were similar to those observed for HIVST. Syphilis self-testing may increase syphilis test uptake among sexual minorities in Zimbabwe and other low- and middle-income countries.

**Key messages:**

- Syphilis self-testing is an empowering, innovative tool that can be used to expand uptake of STI testing among sexual minorities in Zimbabwe.
- Facilitators and barriers for syphilis self-testing are similar to those observed for HIV self-testing in Zimbabwe and other low- and middle-income countries.
- Participants reported high self-test usability and found that self-testing provided increased privacy, convenience and autonomy in comparison to facility-based testing.

## Introduction

In 2016, WHO estimated 19.9 million cases of syphilis worldwide, with the highest prevalence in the WHO African region (1). In the same year, the Global Health Sector Strategy on Sexually Transmitted Infections sets an impact goal to reduce syphilis infections by 90% globally between 2016-2030. As syphilis is often asymptomatic, testing is essential to effectively interrupting transmission and innovative strategies are needed to expand syphilis test uptake (2). Syphilis is more common among men who have sex with men (MSM), with the WHO reporting a median seroprevalence of 6.0% in this group, estimated from 2016-2017 Global AIDS Monitoring data (3). A 2020 biobehavioural survey in Zimbabwe found that 5.1% of Harare MSM had positive treponemal and non-treponemal tests (4). In addition, syphilis and HIV share common sexual risk behaviours and syphilis facilitates HIV transmission, making syphilis co-infection particularly prevalent in HIV-infected MSM (2)(5). As PrEP becomes increasingly available in LMIC, risk behaviours may also change and inadvertently facilitate STI transmission (6). As a result, the WHO strongly recommends routine syphilis screening among MSM (7).

MSM face unique health care challenges because of lack of funding for MSM health, lack of testing, legal and cultural barriers, and stigmatisation, particularly in low- and middle-income countries (LMIC) (8). Stigma associated with same sex relationships may also extend to healthcare facilities and professionals (9). There is also a considerable gap in evidence to guide MSM health programs in many LMICs (10). As a result, despite WHO recommendations, MSM are frequently excluded from syphilis testing services in many LMICs (2).

One way to expand MSM syphilis test uptake is self-testing. Syphilis self-testing is an approach whereby a person performs a rapid syphilis test themselves and interprets the result in private. Self-testing may overcome some of the barriers associated with facility-based testing, promoting early diagnosis, interrupting disease progression, and reducing syphilis transmission (11).

HIVST is recommended by the WHO to expand test uptake among stigmatised key populations (7). A qualitative evidence synthesis found that HIVST empowered people and decreased test-associated stigma (12). Many countries, including Zimbabwe, have policies to support HIVST as an entry point into sexual health services (13). However, there is less evidence supporting syphilis self-testing, despite the known importance of qualitative research in implementing novel diagnostic technologies (12). Syphilis self-testing pilots have shown that it may increase testing frequency by empowering MSM and reducing the impact of structural barriers, but there is no data from sub-Saharan Africa (14)(15). Additionally, in the context of the COVID-19 pandemic, self-testing has become an increasingly important pathway to safely sustain testing when testing facilities are closed or only partially open.

This study aims to understand how syphilis self-testing can create opportunities to test for MSM in Zimbabwe. The purpose of this study was to determine facilitators and barriers for syphilis self-testing and to assess the usability of syphilis self-testing as reported by Zimbabwean men who have sex with men (MSM).

## Methods

A two-phased mixed methods study was conducted among MSM in Zimbabwe. We focused on Harare because of the strong network of MSM community-based organisations in the city. The first phase was prior to syphilis self-testing and the second phase was after syphilis self-testing. The formative data from both phases informed a trial protocol aiming to compare syphilis self-testing to facility-based testing in MSM in Zimbabwe (16).

In phase one, in-depth interviews were conducted amongst MSM and key informants, by trained and experienced researchers from the Pangaea Zimbabwe Aids Trust (PZAT), between March and April 2020. We recruited MSM using snowball sampling (17). Participants needed to meet the following inclusion criteria: 16 years or older, living in Harare, ever had anal or oral sex with another man, born biologically male, and able to provide informed consent. All MSM were referral facilitators, responsible for offering community support to individuals who are harder-to-reach. Key informants were healthcare professionals and were purposively sampled to include providers who had experience with HIV and/or syphilis testing.

Interviews were conducted using a structured guide, lasted approximately 30 minutes and were audio-recorded. The MSM interview guide was developed to explore prior syphilis- and HIV-testing experiences, facilitating and deterring factors, and self-testing intervention preferences. Socio-demographic data were also collected. The key informant interview guide included healthcare provider experiences with HIV and syphilis testing and treatment services, including population served and challenges faced.

Interviews were translated and transcribed by PZAT researchers. Transcripts were then entered into Dedoose 8.3.17. The Framework Method was used to guide our analysis (18). Two codebooks were developed based on an adapted Social Ecological Model to systematically analyse the data, following calculation of the intercoder agreement. Ultimately our conceptual framework included an individual level, a community level and a policy and environment-level (Figure 1) (19). The framework was used to organise deductive and inductive themes emerging from the data, and to create separate analytic memos for MSM and key informant data. The preliminary findings described in these analytic memos were used to refine the pilot trial protocol (MRCZ/A/ 2533).

**Figure 1:**
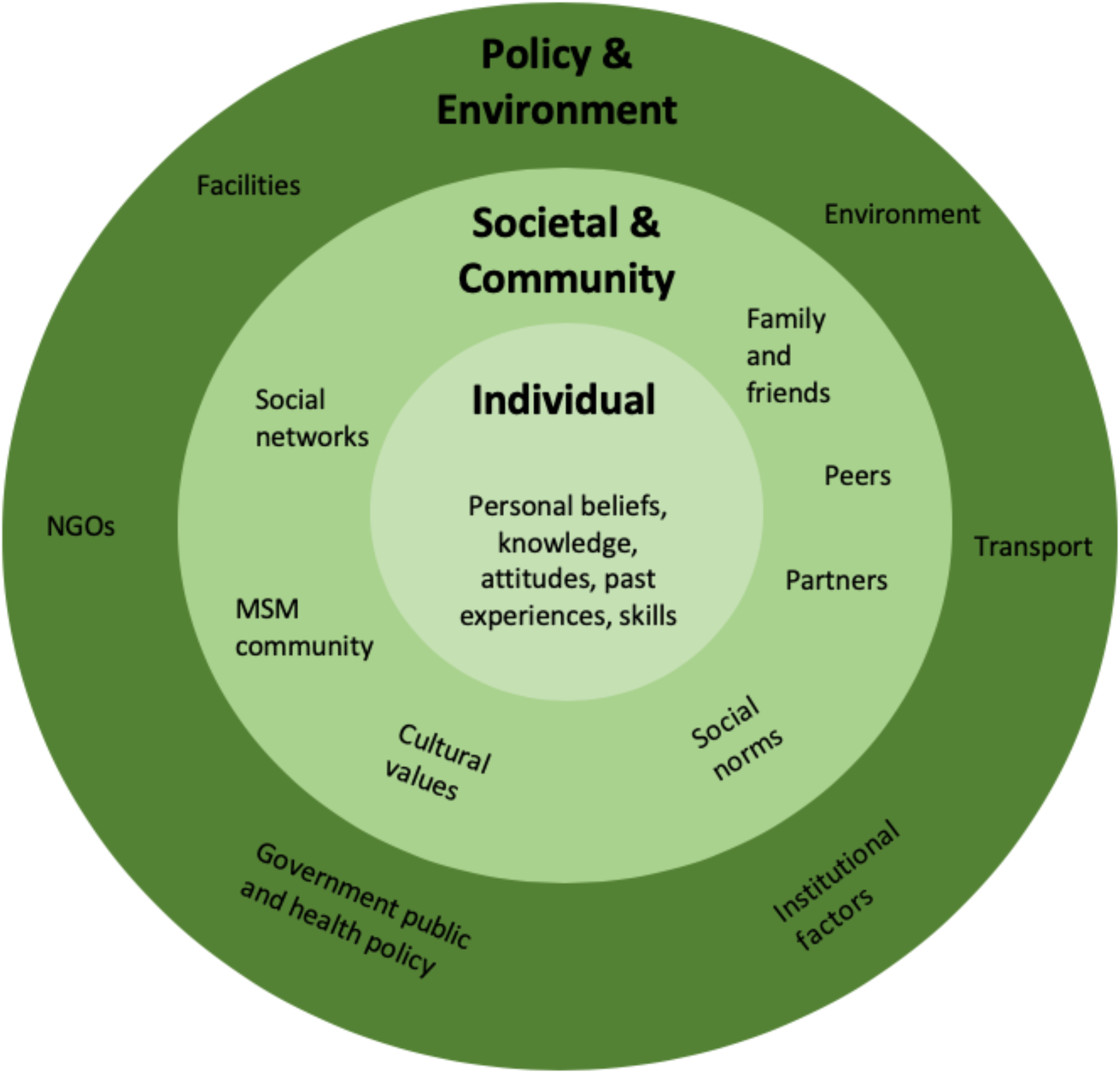
An adapted social-ecological framework of factors influencing test uptake and acceptability of a syphilis self-testing intervention among MSM (19)

In phase two, the syphilis self-test distributed to MSM consisted of a Standard Q Syphilis Ab treponemal blood-based rapid test (SD Biosensor), adapted for individual use and interpretation. Individual lancets and buffer samples were packaged into sealed plastic pouch, together with an individual test device and an infographic, created to explain step-by-step use and appropriate disposal of the kit. An instructional video was created and disseminated to facilitate independent use. Tests were distributed by researchers from PZAT to the same MSM who had completed in-depth interviews in phase one. It emerged that insufficient quantities of buffer were provided in some test kits, impeding successful self-test completion. This was however resolved through community-based distribution of additional buffer samples.

In phase two (August 2020), PZAT researchers interviewed a sample of 16 MSM who successfully completed a syphilis self-test. These interviews were conducted under COVID-19 social distancing measures, as per guidance provided by the Zimbabwe Ministry of Health. Only 16 MSM were interviewed in phase two, as four of the initial cohort of 20 MSM were lost to follow-up. An exit interview guide was developed to qualitatively assess specific facilitators and barriers for syphilis self-testing. Participants also completed a survey to establish quantitative usability of the test, adapted from a HIVST usability index used in South Africa (20). Qualitative data was analysed following the Rapid Assessment Procedures (RAP), a set of methodologies designed for rapid assessment of health-seeking behaviour (21). A RAP matrix was developed using the exit interview guide and Social Ecological Model. Data were then organised into the matrix, by paraphrasing, synthesizing and quoting from participant responses in interview recordings. This allowed us to simultaneously and systematically identify similarities, differences and trends in responses (22). A table illustrating the template that was used by researchers to analyse participant responses regarding the syphilis self-testing kits can be found in Appendix 3.

Ethical clearance was obtained from both Zimbabwe (MRC/A/2533) and London School of Hygiene and Tropical Medicine (Ref: 17848). In line with MRCZ guidance, participants were each compensated for their time. Participants provided informed verbal consent before the start of all interviews. Data was anonymised at the source and participants were given a unique ID.

## Results

Twenty MSM and 10 key informants were recruited for in-depth interviews in phase one. In phase two, the whole cohort was contacted but four MSM were lost to follow-up due to relocation or communication difficulties. Sixteen MSM were therefore invited to conduct the self-test independently and were subsequently interviewed. In phase one, 18 of 20 MSM had previously used HIVST (Table 1). All of these MSM had at least secondary-level education and all but three self-identified as MSM. We observed the following themes in qualitative data: prior STI and HIV testing experiences, both with self-testing and facility-based services; usability of the syphilis self-test and how it compares to HIV self-testing; MSM-specific facilitators and barriers for self-testing.

**Table 1.** Demographic characteristics of in-depth interview participants and exit interviews following the syphilis self-test kit trial

|  | Men who have sex with<br>men<br>n (%) |
| --- | --- |
| **Phase 1 – in-depth interview** |  |
| **Age** |  |
| Mean age in years (range) | 24 (20-33) |
| **Education level** |  |
| Secondary | 7 (35.0) |
| College | 6 (30.0) |
| University | 7 (35.0) |
| **Employment status** |  |
| Student | 7 (35.0) |
| Formal work | 7 (35.0) |
| Informal work | 3 (15.0) |
| Unemployed | 2 (10.0) |
| Other | 1 (5.0) |
| **Sexual orientation** |  |
| Gay (MSM) | 17 (85.0) |
| Heterosexual (MSW) | 2 (10.0) |
| Bisexual (MSM/MSW) | 1 (5.0) |
| **Self-reported disclosure of sexual identity** |  |
| Out to family, friends or doctors | 19 (95.0) |
| Not out | 1 (5.0) |
| **History of HIV self-testing** |  |
| Yes | 18 (90.0) |
| No | 2 (10.0) |
| **Phase 2 – syphilis self-testing exit interview** |  |
| History of syphilis facility-based testing | 8 (50.0) |
| Conducted a syphilis self-test | 16 (80.0) |
| Tested positive for syphilis | 2 (12.5) |
| **Self-reported disclosure of sexual identity** |  |
| In person via community-based organisation | 6 (37.5) |
| Through messaging via WhatsApp | 9 (56.25) |

### Prior HIV and STI testing experiences

In phase one, eighteen MSM had experienced HIV self-testing before using the oral HIV test. Ten participants stated they used HIVST every three to six months. In addition, thirteen of the 18 HIVST-experienced MSM had then attended a facility and were empowered to seek facility-based services. Key informants confirmed that syphilis testing is usually reserved for pregnant women, and only three had received training on how to work with MSM, suggesting MSM are largely neglected by STI services. Some providers recommended syphilis testing should be mandatory for key populations.

### Syphilis self-test usability and comparison with HIVST

Of the 16 participants in phase two, two (12.5%) tested positive for syphilis. Fifteen participants reported the clarity of explanations provided in the infographic and video were instrumental to successful test completion. Overall, MSM reported 89.6% usability for the syphilis self-test on a 0-100 scale. This is described in detail in Table 2. The main challenge with the test kit, reported by 11 of the 16 participants, was the blood draw using the capillary pipette. Participants nonetheless felt this particular challenge was warranted for the test to function. One participant had difficulties extracting the buffer because insufficient quantities were provided. Four participants had to repeat the test, as they did not provide enough blood for the test to show a result.

**Table 2.** Usability index of the syphilis self-test based on a stepwise questionnaire administered in phase two

| Usability Checklist | YES<br>n | NO<br>n | Usability index<br>(%)<sup>‡</sup> |
| --- | --- | --- | --- |
| *Did you find it easy to read/use the information sheet?* | 16 | 0 | 100 |
| *Did you find it easy to watch/use the instructional video?* | 16 | 0 | 100 |
| *Was it difficult for you to remove the kit components from the pack?* | 1 | 15 | 93.8 |
| *Did you verify that the silica gel pouch was yellow, to confirm their test was valid for use?* | 16 | 0 | 100 |
| *Did you remove the yellow shield from the lancet?* | 16 | 0 | 100 |
| *Did you have difficulty lancing (pricking) their finger using the blue lancet?* | 1 | 15 | 93.8 |
| *Did you have difficulty forming a blood droplet?* | 5 | 11 | 68.8 |
| *Were you able to pick up a blood drop up to the black line of the capillary pipette?* | 5 | 11 | 31.2 |
| *Were you able to open the green buffer bottle?* | 16 | 0 | 100 |
| *Were you able to use the pink pipette to pick up the buffer?* | 15 | 1 | 93.8 |
| *Did you drop three drops into the test device well?* | 15 | 1 | 93.8 |
| *Was a control line present on the test device?* | 12 | 4 | 75.0 |
| *Did you trust the self-test result?* | 15 | 1 | 93.8 |
| *Did you quit the process at any point?* | 0 | 16 | 100 |
| *Did you continue the process despite a missed or incorrect step?* | 0 | 16 | 100 |
| Total Usability Index |  |  | 89.6% |

<sup>‡</sup> The usability index (UI) was calculated based on the method used in the HIVST paper from which the index itself was extracted. The original UI is based on WHO literature on Diagnostic Assessment for submission to prequalification. Like in the HIVST study, we tracked all successful steps, in order to quantify a usability index, expressed as a percentage (20)

### Comparing syphilis self-testing to HIVST

Phase two participants felt that the syphilis and HIV self-test kits had many similarities, including the potential for privacy and convenience. The major challenge cited was that syphilis self-testing uses a blood sample whilst most HIVST kits use oral samples. Two MSM reported a preference for HIVST compared to syphilis self-testing because of this issue. However, fifteen (94%) participants felt that they trusted the syphilis test result more because it was blood-based. They also preferred the syphilis self-test because of the clarity of instructions compared to prior HIVST instructional material.

### Self-testing facilitators and barriers

Facilitating and deterring factors for self-testing were categorised into individual, community and structural-level factors (Table 3). Convenience, privacy, and autonomy were the most cited reasons why MSM preferred self-testing over facility-based testing.

**Table 3:** Summary of facilitating and deterring factors influencing MSM testing decision, including quotes from phase one in-depth interviews

| SEM level | Facilitators | Quotes | Barriers | Quotes |
| --- | --- | --- | --- | --- |
| INDIVIDUAL | **Privacy** | *But sometimes you need privacy because not everyone is reliable enough to keep your information with and we are humans (on HIVST, 23yo MSM)* | **Blood sample required** | *I was afraid of being pricked (MSM020, on HIVST)* |
|  | **Autonomy and self-empowerment** | *One, I do the thing on my own when I'm willing to do it. Two, it produces the results that I will see on my own" (on HIVST, MSM age unspecified)* | **Reluctance to test and poor awareness surrounding syphilis** | *At first, I was scared of being positive but my cousin encouraged me to go for HIV tests and told me that if I become positive that will not be the end of life' (on HIVST, 20yo MSM)* |
|  | **User-friendly testing and innovation** | *It's because it's an improvement. Things will be better than [facility-based syphilis testing] where you go there and they take the blood (on syphilis self-testing, 27yo MSM)* |  | *I wouldn't [consider taking a syphilis self-test] because in my mind I have already told myself that I do not have STIs, maybe I would encourage others instead. (on syphilis self-testing, 23yo MSM)* |
|  | **High perceived trust in blood-based tests** | *I feel the blood based one gives a more accurate result. (on HIVST, 21yo MSM)* |  |  |
| COMMUNITY | **Avoidance of social stigma** | *The MSM community is small [...] mostly they will spread rumours that you don't have to date this person because he has syphilis (on syphilis self-testing, 30yo MSM)* | **Stigma at community level over testing within peer groups** | *You would be afraid because the people you live with, if they find out that you are self-testing for HIV, they'll be like "what pushes you?", which means you're practicing something" (on HIVST, 23yo MSM)* |
|  | **High perceived importance of syphilis and peer pressure to test** | *I have a boyfriend of mine who said he has syphilis, so he was treated for syphilis. So I need to see if I also have it (on syphilis self-testing, 24yo MSM)* |  |  |
| STRUCTURAL | **Convenience and improved access** | *No, I was just motivated with the channel of self-testing because sometimes you won't be having any access (on HIVST, 27yo MSM)* | **Indefinite linkage to care and treatment availability** | *We give them kits, but some of them don't come back. [...] they end up knowing your phone number they will end up not answering. So that is the challenge with the self-test (key informant, Kuwadzana Polyclinic)* |
|  | **Avoidance of hostility, stigma, and discrimination from healthcare providers** | *You know sometimes you need to go through a whole lot of protocol to get the test kit and that's what I wouldn't want (MSM005)*<br>*I would not want to go the clinic because some nurses have got an attitude towards people like us because some of them are homophobic. (MSM003,* |  | *Benzathine... rarely. Sometimes it's out of stock and if you refer patients to go and buy outside at pharmacies, that's where there is a challenge (key informant, Hatcliffe clinic)* |
|  | **Time savings** | *I actually did it [...] without having to go anywhere or consult anyone, so yah, in terms of time, in terms of cost it was cost effective." (on HIVST, 25yo MSM)* |  |  |
|  | **Monetary savings** |  |  |  |
|  | **Avoidance of health risk for providers** | *We do not have safety clothes to protect us when we are doing HIV tests [...] and we will be putting our lives in danger especially in this period of COVID-19 (key informant, Malbereign clinic)* |  |  |

### Self-testing facilitators

The following factors were facilitators for both HIVST and syphilis self-testing: privacy, autonomy and empowerment, convenience, user-friendliness, high perceived trust in blood-based tests, avoidance of social and healthcare provider stigma, monetary and time savings, and reduced contact with facility-based services in the COVID-19 context. All MSM participants felt comfortable testing alone and stated they would prefer doing their next test at home, in order to be the first to see their results. In comparison, three participants stated that facility-based testing did not provide adequate levels of privacy. MSM liked that they could conduct their test without the involvement of a healthcare provider and the convenience of it.

MSM highlighted that the lengthy waiting periods for in-facility testing are an important deterring factor. A rapid self-test could contribute to speeding up diagnosis, reducing treatment delay and more efficiently interrupting syphilis transmission. Seven participants in fact mentioned that HIV self-testing empowered them to test more frequently and take control of their sexual health. All phase two participants stated that the blood draw increased their trust in the syphilis self-test. Two MSM noted the blood draw for syphilis facility-based testing is more painful than the self-test, due to the nature of the self-testing lancets provided, and thus would opt for the self-test. Participants explained that they preferred the pressure-activated lancets provided in the study self-test kits, in comparison to the twist-top universal lancets used in-facility.

Participants liked that they were able to avoid being identified at a facility and stigmatized by members of their own community. Additionally, several MSM observed that self-testing prevented hostility from providers or other society members, therefore decreasing test-associated stigma. Key informants in phase one explained they valued self-testing because of the potential to reduce contact with clients, especially in the context of the COVID-19 pandemic.

### Barriers to self-testing

Themes related to barriers included the following: the challenge of self-sampling blood, reluctance to test due to poor awareness, stigma at community-level following at-home testing, indefinite linkage to care and treatment availability. Twelve participants experienced difficulty with the blood draw that they attributed to inexperience. One participant was concerned about the bio-hazard potential with test-kit material disposal. Some MSM mentioned that self-test uptake is jeopardised among the wider community of MSM by poor awareness and the perception that they do not have STIs. MSM also expressed concerns over the fact they could be profiled or stigmatised within their own community following at-home self-testing. Participants reported that they would seek confirmatory testing if trusted information was provided on where to go and what to expect in-facility. These are legitimate concerns that align with phase one qualitative data, which showed that provider discrimination and treatment shortages exist at structural level. Multiple key informants also reported frequent unavailability of the facility-based syphilis tests required for confirmatory testing, as these are reserved for antenatal care.

## Discussion

Our study expands on the limited literature on syphilis self-testing, includes qualitative and quantitative data, and follows MSM prior to and after self-testing. We found that syphilis self-testing was feasible and highly acceptable among MSM in Zimbabwe. The high usability index (89.6%) suggests that syphilis self-testing would be acceptable in this subgroup of MSM. Overall, 12.5% of phase two MSM tested positive for syphilis, a high proportion considering the relatively small number of participants. Participants reported self-testing was a convenient method that provided increased privacy, autonomy and diminished vulnerability in comparison to facility-based testing. The testing challenges associated with the amount of test buffer were transient and were improved by increasing the quantity of buffer provided.

Study findings are consistent with HIV self-testing data in Zimbabwe, as well as syphilis self-testing data from China (23) and the Netherlands (15). Our qualitative data suggested that many of the same facilitators and barriers for syphilis self-testing exist for HIV self-testing. Self-testing is a private and convenient method that is preferred over facility-base testing, especially for higher risk individuals. This is reflected in the large body of evidence that exists for HIV self-testing, which is now well established in Zimbabwe (24). We found that syphilis self-testing was the first ever syphilis test for half of our study participants. This is consistent with data from China suggesting that syphilis self-testing may increase test uptake among MSM (23). Recent data from HIVSTAR in Malawi, Zambia and Zimbabwe also show that HIVST also encourages first-time HIV testing (25).

Our qualitative data suggest that syphilis self-testing can empower MSM to test when, where, and with whom they wish. This is consistent with a global HIVST qualitative literature showing how self-testing gives agency to those who test (12)(26). Existing research also shows self-testing can improve testing frequency (27,28). Providing autonomy, control and creating a culture of testing among vulnerable MSM could potentially help to build trust in the local health system, which is relatively low according to recent evidence (9).

One barrier to syphilis self-testing was the uncertainty of linking to confirmatory testing and treatment within health facilities. Key informants noted that Zimbabwe hospitals have variable access to non-treponemal tests and stock-outs of penicillin occur. While similar concerns existed for HIVST, linkage to care rates have been excellent (26). Poor linkage to syphilis care would impact the capacity for testing to translate into public health benefits for syphilis control. Embedding syphilis self-testing within the HIVST systems could be a way to enhance linkage to care. HIVST has been part of the Zimbabwe National HIV/AIDS Strategic Plan since 2016. The recent large scale HIVSTAR implementation study used community-based distributors, accounting for over 75% of test kit distribution, through Population Services International branches, achieving 50.3% community-level coverage (25). A number of studies in China show successful integration of HIV and syphilis testing services (11).

This study has a number of limitations. Firstly, as a mixed methods study, qualitative results should be interpreted as only an indication of the preferences of the men interviewed. The MSM participants all had at least secondary-level education, were educated about STIs and able to access community-based services. They may therefore be early adopters within the MSM population, more likely to take up health innovations due to heightened awareness and contact with MSM community organisations (29). Most of the interviewees had tried HIVST, which could have also made them familiar with the self-testing method and thus more likely to accept syphilis self-testing. The perspectives of this subset of MSM may be different to those of other, potentially more marginalised MSM in Zimbabwe. For example, subsets of low literacy MSM have had problems implementing HIVST and this may also be the case for syphilis self-testing (30).

This study has implications for research and policy. It has revealed that more research is needed on how we can integrate syphilis self-testing into established networks of HIV self-testing services to facilitate implementation. Syphilis self-testing cannot effectively contribute to interrupting syphilis transmission if facility-based confirmatory testing and treatment is not made accessible to MSM. Clinical trials are needed to assess the effectiveness and risks of syphilis self-testing in practice. From a policy perspective, many of the existing HIVST policies could be expanded to cover syphilis self-testing. Further policy development will help national leadership to embrace syphilis self-testing as a tool for expanding syphilis testing. Improving testing among key populations can reduce the bridging of syphilis into the general population, likely having an impact on the overall prevalence of syphilis, with the potential of reducing mother-to-child transmission.

In conclusion, the findings from this study suggest that syphilis self-testing may decrease user perceived test-associated stigma and empower MSM in an area where same sex relations are condemned. As PrEP is expanded in Zimbabwe and other LMIC settings, leading to a possible shift in sexual risk behaviours, syphilis prevalence may increase. Innovative tools such as syphilis self-testing are needed to expand syphilis test uptake, especially for marginalised populations of MSM.

## Supporting information

1- Phase one in-depth interview guide for MSM

2- Phase one in-depth interview guide for key informants

3- Phase two post-testing in-depth interview guide for MSM

4- Syphilis self-testing infographic distributed as part of the syphilis self - testing kit

## Data Availability

Data are available upon reasonable request : All individual patient data collected that underlie the results reported in this article will be available (text, tables, figures and appendices). Analytic codebooks and consent forms are also available upon request. This data will be available immediately following publication to researchers who provide a methodologically sound proposal. Proposals should be directed to. To gain access, data requestors will need to sign a data access agreement.

## Acknowledgements

The authors acknowledge the support of the Ministry of Health and Child Care, City of Harare and partner organisations who were instrumental in completing this research, including GALZ, Hands of Hope and Zimbabwe Rainbow Community (ZRC). We acknowledge in particular the support provided by Dr Hilda Bara and Ms Anna Machiha. Last but not least, the authors would like to thank all the men who gave their time to participate in both phases of this study in Harare.

## Contributors

JT, MM and IM designed the study. CS, DN, TM and GG analysed the data under the supervision of JT. CS wrote the initial manuscript. All authors were involved in revising the manuscript and approved the final version.

## Patient consent for publication

Not required

## Ethics approval

Ethical approval was granted by the London School of Hygiene and Tropical Medicine (Ref: 17848) and the Medical Research Council of Zimbabwe (MRC/A/2533). All participants provided informed consent.

## Provenance and peer review

Not commissioned; awaiting external peer review.

## Data availability statement – Data are available upon reasonable request

All individual patient data collected that underlie the results reported in this article will be available (text, tables, figures and appendices). Analytic codebooks and consent forms are also available upon request. This data will be available immediately following publication to researchers who provide a methodologically sound proposal. Proposals should be directed to. To gain access, data requestors will need to sign a data access agreement.

