## Supplementary material for "Syphilis self-testing to expand test uptake among men who have sex with men: a theoretically informed mixed methods study in Zimbabwe": 1- Phase one in-depth interview guide for MSM

Supplementary material 1 : Phase one in-depth interview guide for MSM

IDI Guide MSM_Formative Work

PZAT
PANGAEA ZIMBABWE
AIDS TRUST

**In-depth Interview (IDI) Guide for Men having sex with men.**

**Syphilis Self-testing to expand test uptake among men who have sex with men (SST): A pilot RCT in Zimbabwe.**

**About this Study:**
You are invited to take part in a research study that will help us better understand whether syphilis self-testing might be a useful strategy for men having sex with men in Zimbabwe. Your participation in this project will allow us to develop better interventions to promote syphilis self-testing and linkage to care among men having sex with men across Zimbabwe.

**What's Involved?**
If you participate in this study, we will ask you some questions about your sociodemographic information and information about HIV/syphilis related sexual behavior and testing. Your information will all be recorded using an audio recorder and the audio will not have your name on it but a study number and you will not be identified in the interview.

**Study ID:**

IDI Guide MSM_Formative Work

**Objectives**

1. To understand experiences from those who have conducted HIVST.
2. To understand preferred distribution channels for SST.
3. To understand how MSM would want to access the SST.

**Warm-up, knowledge and attitudes for HIV self-testing (HIVST)**

1. Have you ever been tested for HIV? (Y/N)
    a. What has been your experience with HIV testing?

2. Where have you used facility-based HIV testing services? (clinic, hospital, mobile clinic)

3. Are you aware of HIVST? (Y/N)
4. Can you tell us what you know about HIVST?
    - How did you learn about it? (who/where? Peer, community facilitator, facility)
    - Have you ever tested yourself for HIV? (Y/N)
    - Which test did you use? (blood based/ orasure?)

5. Have you ever used HIVST?
    i. If yes,
        - When was the last time you did the HIVST?
        - Do you use it often?
        - How often do you use HIVST?
        - Did you prefer it to other forms of testing? Why?
    ii. If no,
        - Why not?
        - Do you have any concerns with HIVST?

**Personal experiences with HIVST**

6. Can you describe in detail your first time using an HIVST?
    a. How did you obtain it? (peers, community facilitators, facility)
    b. Was this your first time testing for HIV?
        i. If yes,
        - What barriers to facility based testing did HIVST overcome?
        - Did using an HIVST encourage you to later get facility based testing?
        - What was your motivation for taking an HIVST?

        ii. If no,
        - Did HIVST encourage you to later get facility based testing?
    c. What was the context? (Anyone forced you?), why (motivators), when, where
    d. What was your initial reaction to the result? (anxious, sad, suicide)
    e. How did you feel about the accuracy of the result? Do you trust it?
    f. How did you feel about the process? Any difficulties?
    g. Did you tell anyone of your result? (partner, peer, family, health care provider) –
    h. Did you take the test with anyone or alone?
    i. Did you receive any money for returning your result?

7. What motivated you to choose HIVST?
    a. Facilitators? (convenience, privacy, autonomy)
    b. Barriers? (cost, reliability/quality)
8. Concerns before/during/after? (stigma, confidentiality/protection of privacy)

9. In what way has HIVST affected your life? (increase test frequency, condom usage)

**Syphilis self-testing (SST)**

10. Have you ever used facility-based syphilis testing services?
    - Where have you used facility-based syphilis testing?
    - How often?
    - Have you ever tested positive to syphilis? When? Where?
11. Would you consider taking a syphilis self-test?

    If yes,
    - Why? (Facilitators)

    If no,
    - Why not? (Barriers)

    a. Where would you want to access the test? (home, facility, community based organization)
    b. How would you want to access the self-test? (alone, health care provider, peers, community facilitator, partner, family)
    c. Where would you want to take the test? (home, facility)
    d. Would you prefer it to facility based testing? Why?

12. Would you be able to tell anyone about your syphilis self-test result? (partners, friends, family)

13. Would you be able to send your results to a study staff member if you were to take the test at home using a smart phone?

    If yes,
    - Why (have access to a smart phone, have phone credit)

    If no,
    - Why not?

14. How serious a disease do you think syphilis is?
