## Supplementary material for "Syphilis self-testing to expand test uptake among men who have sex with men: a theoretically informed mixed methods study in Zimbabwe": 2- Phase one in-depth interview guide for key informants

### Key Informant Interview (KII) guide

PZAT
PANGAEA ZIMBABWE
AIDS TRUST

**Syphilis Self-testing to expand test uptake among men who have sex with men (SST): A pilot RCT in Zimbabwe.**

**About this Study:**

You are invited to take part in a research study that will help us better understand whether syphilis self-testing might be a useful strategy for men having sex with men in Zimbabwe. Your participation in this project will allow us to develop better interventions to promote syphilis self-testing and linkage to care among men having sex with men across Zimbabwe.

**What's Involved?**

If you participate in this study, we will ask you some questions about your health care work in the area of HIV/syphilis testing and treatment. Your information will all be recorded using an audio recorder and the audio will not have your name on it but a study number and you will not be identified in the interview.

**Study ID:**

#### Objectives

1. To understand health care providers (HCP) experiences with HIVST and linkage to services.
2. To understand HCP experiences with syphilis testing.
3. To understand Community Facilitators (CF) experiences with HIVST services.

#### Demographic Information

1. How long have you been a health care provider?
2. How long have you been at this facility?
3. Which population do you normally serve?

##### HIV Self Testing

4. What has been your experience with facility based HIV testing?
   a. What support services do you offer? (counselling, referrals)
   b. What are some of the challenges that you have faced when delivering HTS services among MSM?
   c. How have you dealt with these challenges?
5. Do you offer HIVST services?
6. Which populations are you encouraging to self-test for HIV? (why?)
7. What has been your experience with HIVST?
   a. What are some of the challenges that you have faced? (probe for HIVST among MSM)
   b. How have you dealt with these challenges?

##### Syphilis self-testing

8. Do you routinely conduct syphilis testing within your facility?
9. Who do you normally target for syphilis testing?
10. Do you routinely see MSM within your facility?
    If yes,
    - What services do they normally come for?
    - What has been your experience with syphilis testing at this facility?
11. Do you routinely offer treatment to those who test positive to syphilis?
12. What has been your experience with syphilis screening and treatment? (screening, drug availability, incomplete treatment)
13. Are there any challenges that you have observed in relation to syphilis screening and treatment among MSM?
14. How prevalent is syphilis among MSM?
15. Resources permitting, what would be your recommendations for syphilis screening and treatment among the MSM population?
