## Supplementary material for "Syphilis self-testing to expand test uptake among men who have sex with men: a theoretically informed mixed methods study in Zimbabwe": 3- Phase two post-testing in-depth interview guide for MSM

PZAT
PANGAEA ZIMBABWE
AIDS TRUST

### Exit interview guide for MSM field test of syphilis self-testing kits

**Syphilis Self-testing to expand test uptake among men who have sex with men (SST): A pilot RCT in Zimbabwe.**

**About this Study:**
You are invited to take part in a research study that will help us better understand whether syphilis self-testing might be a useful strategy for men having sex with men in Zimbabwe. Your participation in this project will allow us to develop better interventions to promote syphilis self-testing and linkage to care among men having sex with men across Zimbabwe.

**What's Involved?**
If you participate in this study, we will ask you some questions about your sociodemographic information. We will also ask you to complete a syphilis self-test, and give feedback on your experience. Your information will all be recorded using an audio recorder and the audio will not have your name on it but a study number and you will not be identified in the interview.

***Is this all clear to you and are you happy for us to interview you today?***

**A) Participant and interviewer information**

**Patient ID:** *E001 to E020*
(use E001 for the exit interview corresponding to the IDI with MSM001, E002 for MSM002, etc.)
**Interviewer name:**
**Interview date:**
**Interview venue:**

**B) I would like to ask a few questions about your syphilis self-test today, and what you thought about it.**

**I. General usability**

Q1. How was your experience with your syphilis self-test kit today?

Q2. Did you face any challenges or find any specific parts of the self-test difficult to perform?

Probe: if the participant expresses difficulty with the finger prick:
Q2.1 'Do you feel that drawing blood is a challenge you are willing to accept for this test to work?'

PZAT
PANGAEA ZIMBABWE
AIDS TRUST

### II. Usability index

https://journals.plos.org/plosone/article?id=10.1371/journal.pone.0227198

| **STANDARD Q test usability index** | YES | NO |
|---|---|---|
| Did you find it easy to read/use the information sheet? | | |
| Did you find it easy to watch/use the instructional video? | | |
| Was it difficult for you to remove the kit components from the pack? | | |
| Did you verify that the silica gel pouch was yellow, to confirm their test was valid for use? | | |
| Did you remove the yellow shield from the lancet? | | |
| Did you have difficulty lancing (pricking) their finger using the blue lancet? | | |
| Did you have difficulty forming a blood droplet? | | |
| Were you able to pick up a blood drop up to the black line of the capillary pipette? | | |
| Were you able to open the green buffer bottle? | | |
| Were you able to use the pink pipette to pick up the buffer? | | |
| Did you drop three drops into the test device well? | | |
| Was a control line present on the test device? | | |
| Did you trust the self-test result? | | |
| Did you quit the process at any point? | | |
| Did you continue the process despite a missed or incorrect step? | | |

### III. Instructions

Q3. Is there anything you would change in the written and pictorial instructions?

Q4. Is there anything you would change in the self-testing video?

Q5. Did you rely more on the video or the written instructions for the self-test?

### IV. Attitude towards syphilis self-testing

Q6. a) Was this your first time testing for syphilis?
b) If not, please detail your experience with other forms of testing

Q7. How do you think this self-test compares to facility-based testing for syphilis? And why?

Q8. Are there any particular factors that make syphilis self-testing better than facility testing?

Q9. Are there any particular factors that make syphilis self-testing worse than facility testing?

Q10. If you had the opportunity to test for syphilis again, would you choose a self-test? Why?

Q11. Would you recommend this syphilis self-test to a friend or family member? And why?

Q12. Do you have any particular concerns regarding this syphilis self-test?

Q13. What would you change about this self-test?

PZAT
PANGAEA ZIMBABWE AIDS TRUST

### V. Participant satisfaction with the syphilis self-test

Q14. a) Did you feel comfortable completing this self-test alone at home? Why?
b) If not, what were the reasons for which you felt unsafe?

Q15. Did you trust the syphilis self-test result and why?

Q16. If your result had been ***positive***, how do you think you would have reacted? [***If negative****, ask how participant would have reacted*]

a) Would you have been able to send the result to us?
b) Would you be able to go to a health care facility to get treated? [*Ask only if positive*]
c) What kind of information and support would you have wanted in the event that you tested positive? [*Ask only if positive*]

d) . Would you disclose your test result to your sexual partner(s)?
    - If the test was negative?
    - If it was positive?

### VI. Comparison to HIVST (questions reflect the MSM IDIs)

Q17. Have you used a HIV self-test before?
Q18.
a) If so, how does the syphilis self-test compare to HIV self-testing?

b) What was similar about syphilis self-testing and HIV self-testing?
c) What was different about SST and HIVST?
d) Did you find that you trusted the syphilis test result more because it is a blood-based test?

Q19. Do you have a preference between the syphilis self-test and the HIV self-test and why?

### VII. SST access preferences

Q20.
a) Now that you have tried the syphilis self-test, where do you think you would prefer to collect or receive it? **Prompts**: in the post at home, through a CBO, through a health facility
b) Would you prefer to collect the self-test alone or with the help of someone?

Q21. Where would you prefer to conduct your next syphilis self- test?

***Prompts:***

a) Would you feel comfortable completing a syphilis self-test kit at home and why?
b) **OR** Would you prefer to do the self-test at a facility and why?

Q22.

a) In the context of our research study, would you find it acceptable to send a photo of your completed test kit to one of our research co-ordinators for confirmation? This process would be anonymous.
b) Do you think most men in your community would have access to a smartphone to complete this confirmation process?
c) **If not**, which do you think is the best way to complete a visual confirmation of your self-test result with study coordinators?

Q23. How important do you feel it is to test for syphilis?

Q24. How do you think the availability of syphilis self-test kits would change your life?
