## Supplementary material for "Syphilis self-testing to expand test uptake among men who have sex with men: a theoretically informed mixed methods study in Zimbabwe": 4- Syphilis self-testing infographic distributed as part of the syphilis self - testing kit

### SYPHILIS

Syphilis is a sexually transmitted infection (STI) caused by a bacteria. It is important to test yourself for syphilis if you think you may have been at risk of getting infected. If it is left untreated, it can lead to serious consequences for your health.

#### What are the signs and symptoms?

The symptoms are often unnoticeable and you could be infected without knowing it. Symptoms include:
- sores/ulcers on the penis, anus and around the mouth
- blotchy red rashes on the hands or feet
- skin growths around the anus (similar to genital warts)
- white patches in the mouth
- tiredness, headaches, joint pain, high temperature
- swollen glands in the neck, groin or armpits.

#### Why test myself?

It is difficult to diagnose syphilis from only symptoms because these are usually not specific. In fact, a lot of cases don't show any symptoms at all. Testing for the disease is important because it means you can get treated quickly, to avoid developing complications and transmitting syphilis to your partner.

Being infected with syphilis can also make it easier for HIV to be transmitted through sex. Testing and treating syphilis can therefore help to reduce your risk of getting HIV as well.

#### Why men who have sex with men (MSM)?

Syphilis is a global public health problem, particularly among MSM in urban settings, where more and more infections are It has been found that MSM are at higher risk of getting syphilis. This could be because less men are ensuring safe sex practices and using condoms.
In MSM that are HIV+, syphilis has been reported to present differently and progress more rapidly than if HIV-. Syphilis complications involving the brain are also more common in HIV+ patients.

#### How can I prevent this disease ?

By practising **safer sex:** use a male condom consistently during oral and anal sex; avoid sharing sex toys with people. Be mindful of who you are having sex with and if they are at risk of being infected with syphilis. Having more than one sexual partner and concurrent partnerships also increase the risk of getting syphilis.

#### How is it treated?

Syphilis will not go away on its own.
If it is caught early, it can be treated with 1 injection of benzathine penicillin G or with a longer course of antibiotics.

You should avoid any sexual activity or close sexual contact until 2 weeks after treatment for syphilis finishes. Treatment will also reduce the risk of spreading the infection to others.

#### The disease

In the early stages of infection, the symptoms of syphilis are usually mild and non severe.
If untreated, syphilis infections can progress to a severe stage where patients experience severe medical complications that affect the heart and the brain. There can also be severe skin and/or bone damage and eye problems.
Ultimately, untreated syphilis can lead to death.

#### How is it transmitted?

It is usually caught by having sex or being in close sexual contact with an infected person.
It can also be transmitted from a mother to her child during pregnancy and when giving birth.
You can catch syphilis more than once, even if you have been treated for it before.

#### Where can I get help?

If you think you could have been infected, it is important to seek care as soon as possible. The first step should be to test yourself and to get treated.
There are local facilities that offer these services:
- *Fill with the partner health facilities*

PZAT PANGAEA ZIMBABWE AIDS TRUST

LONDON SCHOOL of HYGIENE &TROPICAL MEDICINE

### SYPHILIS SELF TESTING

#### When should I consider getting myself tested?

You should get tested for syphilis if:
- you have had unprotected sex with one or multiple partners.
- you are worried you might have it.
- you have symptoms of syphilis.
- a sexual partner has been diagnosed with syphilis

The test we offer is the STANDARD Q Syphilis Ab Rapid Diagnostic Test. It will test your blood for syphilis. The test can be carried out in the comfort of your own home

#### Why should I use a self-test?

Self-testing makes it easier for people to know their status:
- it is an easy test you can do whenever and wherever you want.
- There is no need to wait long periods of time : it is a rapid test that only takes 10 minutes.
- you do not have to attend an STI health service : it is anonymous and private.

#### 12 easy steps

**First, watch the self-test video with the QR code below and/or read the instructions from beginning to end.**

1. Verify the test expiry date on the back of the packet.
2. Open the pack. Lay out the test device, capillary pipette, pink 'pipette', green 'buffer' tube, blue lancet, alcohol swab on a clean table. You can also use a phone timer.
3. Check the silica pack: if it is yellow, the test is valid for use. If the silica pack is green, you must not use this test device: please collect a new test pack.
4. Wash your hands. Then use the alcohol wipe to swipe 1 finger clean. Let air dry!
5. Use the blue lancet to prick the finger you cleaned. Use the yellow protective cap to shield the needle. Then dispose of the lancet in the empty test packet.
6. Squeeze your finger to encourage blood flow. Use the capillary pipette to pick up a drop of blood from your finger, it should exactly come up to the black line on the pipette.
7. Squeeze the drop of blood from the pipette into the square well of the test device. Dispose of the pipette in the empty test packet.
8. Add 3 drops of the green buffer into the well where you put the blood. Keep the test device flat on the table.
9. Wait for 5 minutes to read the result. Do not wait longer than 20 minutes to read the result.
10. Interpret the test results
11. Take a photo of the test result
12. Send a photo of the test result along with your patient ID number to your study coordinator.

1

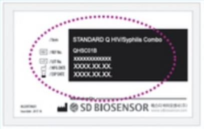


2

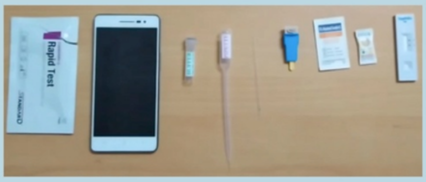


3

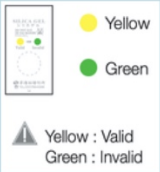


4

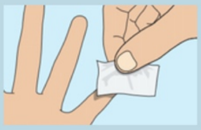

5

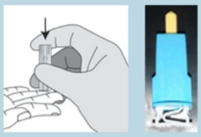

6

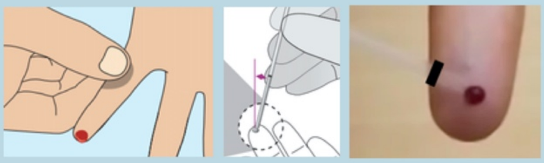

7

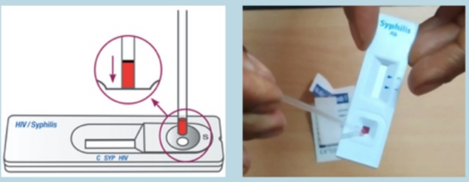


8

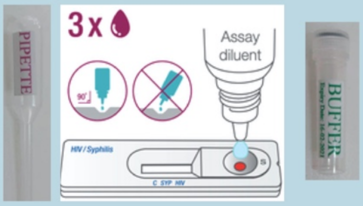


9

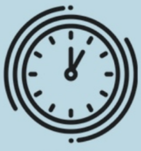

5 min

10

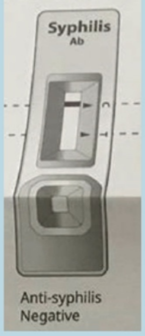


<- Negative: 1 line on 'C' ONLY

Positive -> 2 lines on 'C' AND 'T'

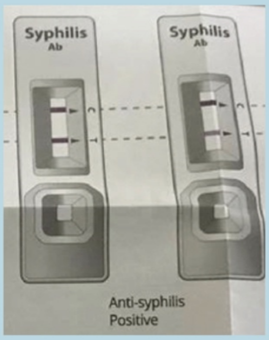


#### How to interpret the test

After 5 minutes the results should have appeared.
The control line 'C' should appear for all results. If the 'C' line does not appear, you will need to complete a new test.
If you obtain a 'C' line only, you are negative. If you obtain a 'C' and a 'T' line, even if it is faint, you are syphilis positive.
If you read that you are positive for syphilis, please go into a facility for a confirmation test and see a doctor as soon as possible for treatment. Please also avoid any sexual relations to avoid transmitting the disease.

SCAN THE QR CODE TO ACCESS A VIDEO ON HOW TO USE THE SELF-TEST.

**Remember that self-test kits are not 100% reliable. If you are worried about anything, get advice from a health professional. If you test positive, contact a health professional to get the medical and emotional support you need.**

### SST FREQUENTLY ASKED QUESTIONS

#### How accurate is the syphilis self-test?

The syphilis self-test is a highly accurate self-test, similar to HIV self-tests. If you follow the instructions carefully and complete the test properly, the test is 100% likely to detect syphilis, according to the test manufacturer. This is a highly sensitive testing instrument and healthcare professionals trust it to diagnose syphilis.

#### I am worried about the finger pricking and I do not like the sight of blood.

It is necessary for this self-test to prick your finger to obtain a few drops of your blood. Finger pricking will provoke a slightly uncomfortable pinch on your finger that will only last a second. The retractable lancet that is provided in the kit makes it easy for you to do this.

This slight pain is a downside to the test, however, it is necessary to complete the self-test properly. Finding out your syphilis status, and getting treatment if you need it, will ensure that you avoid the severe complications of syphilis.

#### How long does it take to complete the self-test?

The syphilis self-test is a relatively short test. The time it takes to complete the test is about 10 minutes, including 5 minutes for the test result to develop.

#### Is it <u>safe</u> for me to complete the test alone at home?

Testing for syphilis can be a stressful experience and it is a natural reaction to be anxious about it. Syphilis is an easily treatable disease so you should not be worried about your diagnosis: the treatment is straightforward and fast acting. There are also counselling services available for you following confirmatory testing and syphilis treatment.

#### I am worried about confidentiality as I want to keep my results private.

We assure you that maintaining confidentiality is among the most important aspects of this study. All self-test results are anonymised straightaway. Our professional research staff is highly qualified and experienced in maintaining the privacy of study participants. The study has gone through extensive ethical review to ensure that all research data remains confidential.

#### Should I tell my friends and family about my result?

We understand that there is stigma attached to a positive STI result. However, we want to reassure you that syphilis is not a serious disease, as it is  easily treatable with a single dose of antibiotics. Early diagnosis and treatment is very beneficial to you and to your sexual partner(s). Although this may be a challenge, we also encourage you to disclose your result to your sexual partner(s). This is because if they are infected with syphilis, and you have completed your testing and treatment, they could easily re-infect you. The whole process would then have been for nothing. We highly recommend that you and your partner regularly test for STIs to avoid re-infection.

#### Do I need to get a second confirmatory test at a health facility?

You must only seek a second confirmatory test in the event that you read a positive result on your self-test. This confirmatory test can be done at one of our partner health facilities. However, please remember that if your self-test result is negative, you can be reassured that you are not infected with syphilis. With a negative self-test, you do not need a confirmatory test.

#### I am worried that the test kit will not work if I store it at home.

This self-test kit is very stable and is completely safe to use outside of health facilities. The self-test is contained in a sealed pack that can be stored in your own home and that will not lose its usability, even if it is outside of a health facility.
